# Policy-Relevant Causal Approach to Assessing the Impact of Occupational Heat and Airborne Particulate Matter Exposure on Acute Kidney Function in Guatemalan Sugarcane Workers

**DOI:** 10.64898/2026.03.20.26348712

**Authors:** Amy Dye-Robinson, Kevin P Josey, Diana Jaramillo, Miranda Dally, Lyndsay Krisher, Jaime Butler-Dawson, Karely Villarreal Hernandez, Alex Cruz, Daniel Pilloni, John L Adgate, Joshua Schaeffer, Richard J Johnson, Michel Chonchol, Lee S Newman

## Abstract

**Background:** Chronic Kidney Disease of unknown etiology is a growing health concern in low- and middle-income countries. Recurrent acute kidney injury has been implicated in the mechanistic pathway to chronic disease. While occupational heat stress is recognized as a potential contributor to kidney dysfunction among agricultural workers, the causal relationship between heat stress, core body temperature (Tc), and kidney function remains unclear.

**Methods:** This was an observational study conducted in Guatemala following 148 male sugarcane workers across two six month harvests. Heat stress was measured using heat index (HI) and Tc with ingestible telemetric temperature pills. Particulate matter (PM) exposure was measured using personal breathing-zone samplers with a size-selective inlet for particles below 5 µm (PM5). Kidney function was measured as the change in estimated glomerular filtration rate (eGFR) from pre- to post-shift. Two doubly-robust analyses were applied: exposure-response curves that describe kidney function across the observed range of each exposure, and modified exposure policies that estimate the change in kidney function under realistic reductions in HI, Tc, and PM.

**Results:** The average daily HI was 37.4 °C (SD: 2.0) with an average Tc increase of 1.16 °C (SD: 0.48) per shift. Both HI and Tc were associated with declines in eGFR across the work shift. At an HI of 34 °C, the estimated average eGFR decline was about 2.5 mL/min/1.73 m² and was not statistically significant, while at 40 °C the decline was about 10 mL/min/1.73 m² and was statistically significant. A modeled policy reducing HI exposure by 5% improved eGFR change by 2.94 mL/min/1.73 m² (95% CI: 0.96, 4.91). No statistically significant association between PM exposure and eGFR decline was detected.

**Conclusion:** Reducing workday heat exposure may mitigate acute kidney function decline. These findings support the development of policy interventions aimed at reducing external heat exposure and internal heat strain to protect kidney health. More research is needed to investigate the potential contribution of other environmental factors, including PM exposure.

## 1. Introduction

Chronic Kidney Disease of unknown etiology (CKDu) is an urgent public health concern, particularly in low- and middle-income countries (LMICs), where chronic diseases are increasingly prevalent among agricultural workers^1 2^. In several regions, notably Latin America and South Asia, chronic kidney disease (CKD) has become one of the leading causes of hospitalization and death, driven in part by the rise of CKDu^3^. Unlike traditional CKD, the disease formation of CKDu typically occurs in the absence of classic risk factors like hypertension, diabetes, or known glomerular disease^4–6^. It remains unclear what cause(s) or interacting risks exist for CKDu, although association studies often implicate working outdoors in extreme heat that can result in acute kidney injury^7^. Evidence from mechanistic and epidemiological studies point to recurrent kidney injury contributing to the development of CKDu^8–10^. Sugarcane harvesters may be susceptible to daily, recurrent AKI during the harvest season^11–12^. Daily reductions in blood flow during intense labor in the heat can cause ischemic damage in the renal tubules leaving them vulnerable to other nephrotoxicants^13^.

Indeed, heat stress is a major occupational health risk worldwide, particularly for agricultural workers who often perform strenuous physical tasks in high-temperature environments^11,14–15^. Prolonged heat stress conditions can trigger heat strain, the body’s physiological response to heat stress measured by core body temperature (Tc), which can escalate to heat-related illnesses, injuries, or even fatalities if preventive measures are not taken^16^. Excessive heat strain can lead to dangerous Tc, and without proper hydration, rest breaks, and cooling, can cause heat-related illnesses and even death.

The human body’s optimal Tc is between 36.1°C and 37.2°C^17–18^. When Tc starts to rise above 37°C, a worker can experience symptoms of heat exhaustion including fatigue, dizziness, and reduced cognitive performance^19–20^. If Tc exceeds 40.6°C, the risk of heatstroke increases sharply, which can result in multi-organ failure, permanent damage to the brain or kidneys, or death^21^. Maintaining Tc within safe limits is necessary for preventing heat-related illness and protecting worker health^22^.

While previous studies have linked CKDu with working and living in hot environments, they also suggest the disease may result from recurrent heat stress, dehydration, intense physical labor, and acute kidney injury^11,23–25^. The design of these studies limits their ability to implicate heat as a causal factor in the etiology of CKDu. Few systematic studies have been conducted in field settings where workers are at high risk for CKDu. Most field studies rely on ambient temperature and humidity as proxies for heat strain^26^, rather than directly measuring the body’s physiological response to heat (i.e., Tc). As a result, prevention strategies for heat-related illnesses and associated chronic diseases, such as CKDu, often depend on extrapolation from laboratory settings, indirect indicators of thermal stress, and assumptions regarding causality and the expected benefit of health interventions ^27^. To add further complexity, previous work has demonstrated that workers who perform intense labor under hot conditions experience concomitant environmental exposures such as airborne particulate matter (PM), including airborne silicates^28–29^. Recent lines of evidence have suggested an association between airborne PM and silica exposure and CKDu^28,30–34^.

Epidemiological studies examining environmental risk factors often focus on identifying associations, limiting their practical utility for guiding prevention efforts or informing policy. Many conclude that more research is needed to establish causal links between exposures and health outcomes. To address these gaps, causal inference methods have been developed to more directly estimate the expected effects of exposures on health outcomes, and to provide results relevant to decision making and policy development. Recent work has applied these methods to establish causal relationships between exposures and health outcomes such as cancer^35^, heart disease^36^, and mortality^37^.

Field-based research on CKDu has the potential to lend empirical evidence that can clarify the causal role of heat exposure and the effectiveness of potential mitigation strategies. Despite major recommendations emerging in the U.S. and globally, there is little data demonstrating the effectiveness of interventions aimed at preventing CKDu. Recent intervention studies have shown that hydration, rest and shade programs often promoted in workplaces can reduce cross-shift AKI and help preserve kidney function among heat-exposed agricultural workers^11,38^. Meanwhile, few studies have examined whether these work shift-level improvements translate into reduced long-term risk for kidney function decline and CKDu^39–40^.

As heat-related illnesses continue to gain attention in occupational settings, applying causal methods can strengthen the evidence base supporting those interventions. This is especially true in occupational field-based settings where modified exposure policies (MEPs), also referred to as modified treatment policies (MTPs), offer a valuable framework for causal inference. By modeling policies as realistic changes to heat exposure, this framework estimates the potential health benefit of interventions a worksite could adopt, such as rescheduling work to cooler hours, enforcing rest-shade-hydration cycles, or lowering the metabolic heat of cutting through work-rate or mechanization changes. In plain terms, this approach estimates how much kidney function would change if every worker’s exposure were lowered by a realistic amount rather than by an unrealistic all-or-nothing change.

In the present study, we apply doubly-robust estimators to empirical data collected under workplace conditions associated with heat-related illness, acute kidney injury, and CKDu, and we estimate two complementary quantities. The exposure-response curve (ERC) describes how acute changes in kidney function vary across the observed range of each heat metric. The modified exposure policy (MEP) estimates the change in kidney function under a realistic, partial reduction in exposure, a quantity that maps onto interventions a worksite could implement. The two analyses are estimated separately but share the same cluster-robust machinery, so the curve and the policy estimates are consistent with one another. Nested within an ongoing research collaboration focused on agricultural workers in Guatemala, this study provides insight into the causal relationship between heat exposure, Tc, PM, and acute kidney function, and it shows how each estimand can guide the design and evaluation of workplace interventions that reduce modifiable environmental exposures.

## 2. Methods

### 2.1. Study Population and Design

This analysis is part of the Chronic Kidney Disease, Heat, and Air Pollution Study (CHAP I), a community-engaged investigation of the effects of air pollution and heat on kidney function. Participants include male sugarcane workers recruited from a large agribusiness in southwest Guatemala during the harvest years of 2021–2022 and 2022– 2023. Participants were eligible if they were aged 18 years or older and actively employed as sugarcane harvesters during the study period. The harvesters at this worksite were exclusively male, so enrollment was limited to men; women in this workforce hold different roles and participated in a different study. As part of the worksite health program, applicants with reduced kidney function are not hired as harvesters for the season. Male harvesters from 12 working groups (“turmas”) were prospectively enrolled in this longitudinal observational study in November, prior to the start of the six-month harvest season, and after providing informed consent to participate. For this study, participants from the two harvest seasons were pooled into one cohort. The two harvest seasons showed only modest differences in ambient heat and cross-shift eGFR change and were pooled to improve statistical precision. The harvest season, which occurs from November to April during Guatemala’s dry season, involves six-day work weeks of physically demanding labor to harvest sugarcane.

Sugarcane harvesting practices have been previously described elsewhere^11,41^. Briefly, sugarcane cutting entails heavy to very heavy physical strain or metabolic rate, as workers progress through fields of recently burned sugarcane, using machetes to trim, cut, and pile rows of sugarcane for approximately eight hours daily. On average, workers self-reported cutting about five tons of sugarcane per day, while company records indicated a somewhat higher amount (about 6.5 tons); we used the company-reported value as the workload measure. A typical workday begins at 7:00 AM and ends at 5:00 PM, with a one-hour lunch break at noon and three 20-minute rest breaks. On particularly hot days, workers may end their shift earlier than 5:00 PM, but they often cut the same amount of sugarcane as full-length days. Shaded areas, usually located near the buses that transport workers to the fields, are provided during breaks. Workers are supplied with fresh potable water, packets of electrolyte solution, and education on the importance of hydration, rest, shade, and symptoms of heat stress and heat illness, as previously described^42–43^.

This study was approved by the Colorado Multiple Institutional Review Board (COMIRB #20–0509) in the United States and ZUGUEME Comité de Ética Independiente in Guatemala.

### 2.2. Data Collection

Survey data, blood and urine samples, and clinical measures were collected four times during the harvest season: at baseline (pre-harvest) and three subsequent time points. During baseline, an initial survey recorded demographic information, health history, behavioral risk factors (e.g., alcohol use, smoking, prior work experience), home environment conditions (e.g., home water sources), and personal health factors (e.g., weight, blood pressure). During the three subsequent time points, survey data were collected on behavioral information including use of non-steroidal anti-inflammatory (NSAID) medications and the number of rest-breaks the worker took during the work shift. Clinical measures included height, weight, and blood pressure. Data regarding the amount of sugarcane cut by each worker was collected by the business and shared with the research team. Workers also self-reported the amount of cane they cut each study day. The time the workers started and stopped working on data collection days was provided by the company and used to calculate the length of the work shift. Heat Index (HI) was assessed using a heat stress monitor, collected continuously in the sugarcane fields with the Kestrel 5400 (Kestrel Instruments, Boothwyn, PA). Core body temperature (Tc) was measured using an ingestible telemetric pill (Bodycap e-Celsius Performance®, Caen, France).

Measurements were collected throughout the entire work shift at three harvest season time points. Time points were approximately two months apart from each other and were selected to capture changes in clinical markers and environmental exposures as the harvest season progressed.

Pre- and post-shift blood samples were collected immediately before and after work by venipuncture and were analyzed by the accredited *Laboratorio Clínico y de Investigación de la Fundación para la salud integral de los Guatemaltecos* (Clinical and Research

Laboratory of the Foundation for the Integral Health of Guatemalans). Creatine kinase was analyzed to evaluate muscle injury, and osmolality was assessed to measure hydration status. Blood creatinine was measured using an iSTAT point-of-care device (Abbott, Princeton, NY, USA) in order to calculate estimated glomerular filtration rate (eGFR) and assess kidney function; the iSTAT has been shown to have high agreement with traditional lab measurements^44^. eGFR was calculated using the 2021 CKD-EPI creatinine equation without a race coefficient, which has been supported for use in this population^45–46^. The outcome of this study was the absolute change in eGFR from the start of the work shift to the end of the work shift.

#### 2.2.1. Core Body Temperature

The telemetric pill was ingested by the participants before the start of their work shift, then collected data continuously throughout the work shift at a sampling rate of 2 minutes. At the end of the work shift, the data were downloaded using the manufacturer’s eViewer Performance monitor (Version 6.1.0). To ensure data quality, the first hour of measurements was excluded during cleaning, because readings in this period reflect sensor equilibration after ingestion and are unreliable. Tc data cleaning methods are described elsewhere^47^. Tc was quantified using two methods: average Tc during the workday and the maximum change in Tc during the workday, which have been shown to be predictive of kidney function when compared to other measures^48^. The maximum change in Tc was calculated by taking the maximum temperature minus the average of the first 60 minutes of the cleaned workday data. This calculated change is referred to as the change in Tc throughout the rest of the paper.

#### 2.2.2. Environmental Measurements

Field-level ambient temperature and relative humidity were measured using a HI monitor (Kestrel 5400, Boothwyn, PA, USA) placed in the same fields where participants worked on clinical and Tc data collection days. HI is a combined measure of air temperature and relative humidity and was calculated according to the National Weather Service formula^49^ by the monitor. HI was quantified as the average HI during the workday for each study day. For this study, HI was used instead of wet-bulb globe temperature (WBGT). A valid WBGT requires the globe (radiant) temperature, and this metric had substantial missingness and quality problems in the field.

Personal breathing-zone measurements of particulate matter <5 microns (PM5) were collected for each worker using a size-selective inlet, following methods described in detail in Adgate et al.^28^. Full-shift time-weighted PM5 exposure (in µg/m^3^) was estimated by dividing total gravimetric filter mass by sampling time and flow rate.

### 2.3. Statistical Methods

Means and standard deviations (SD) for continuous variables and frequencies with percentages for categorical variables are reported in Table 1. The primary outcome was the absolute change in eGFR from the start of the work shift to the end of the work shift (post-shift minus pre-shift) with a negative change representing a decline in eGFR from pre- to post-shift. Three heat metrics were assessed: average ambient HI, change in Tc, and average Tc.

**Table 1.**
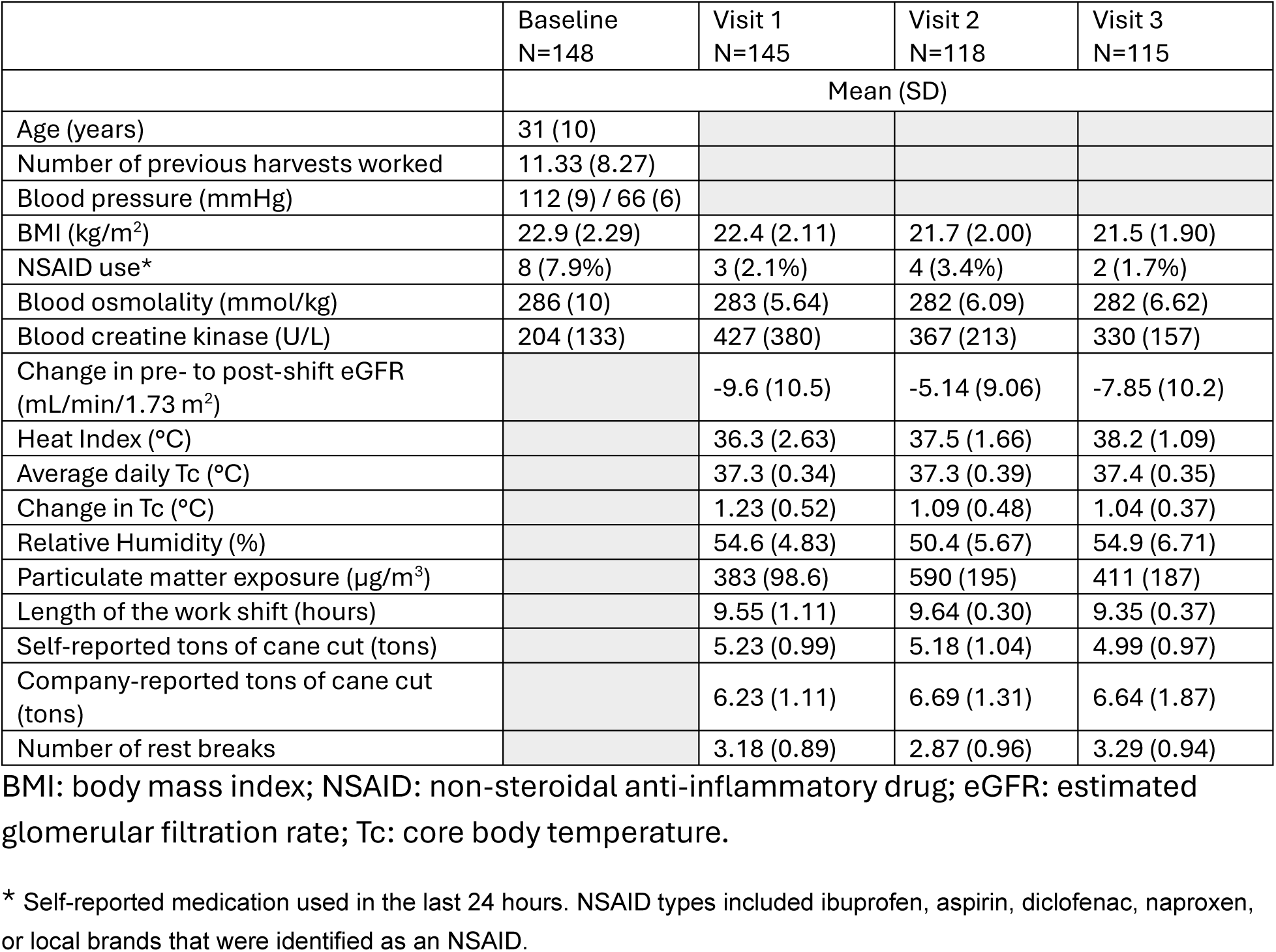
Demographic characteristics of study population.

|  | Baseline<br>N=148 | Visit 1<br>N=145 | Visit 2<br>N=118 | Visit 3<br>N=115 |
| --- | --- | --- | --- | --- |
|  | Mean (SD) |  |  |  |
| Age (years) | 31 (10) |  |  |  |
| Number of previous harvests worked | 11.33 (8.27) |  |  |  |
| Blood pressure (mmHg) | 112 (9) / 66 (6) |  |  |  |
| BMI (kg/m<sup>2</sup>) | 22.9 (2.29) | 22.4 (2.11) | 21.7 (2.00) | 21.5 (1.90) |
| NSAID use* | 8 (7.9%) | 3 (2.1%) | 4 (3.4%) | 2 (1.7%) |
| Blood osmolality (mmol/kg) | 286 (10) | 283 (5.64) | 282 (6.09) | 282 (6.62) |
| Blood creatine kinase (U/L) | 204 (133) | 427 (380) | 367 (213) | 330 (157) |
| Change in pre- to post-shift eGFR (mL/min/1.73 m<sup>2</sup>) |  | -9.6 (10.5) | -5.14 (9.06) | -7.85 (10.2) |
| Heat Index (°C) |  | 36.3 (2.63) | 37.5 (1.66) | 38.2 (1.09) |
| Average daily Tc (°C) |  | 37.3 (0.34) | 37.3 (0.39) | 37.4 (0.35) |
| Change in Tc (°C) |  | 1.23 (0.52) | 1.09 (0.48) | 1.04 (0.37) |
| Relative Humidity (%) |  | 54.6 (4.83) | 50.4 (5.67) | 54.9 (6.71) |
| Particulate matter exposure (µg/m<sup>3</sup>) |  | 383 (98.6) | 590 (195) | 411 (187) |
| Length of the work shift (hours) |  | 9.55 (1.11) | 9.64 (0.30) | 9.35 (0.37) |
| Self-reported tons of cane cut (tons) |  | 5.23 (0.99) | 5.18 (1.04) | 4.99 (0.97) |
| Company-reported tons of cane cut (tons) |  | 6.23 (1.11) | 6.69 (1.31) | 6.64 (1.87) |
| Number of rest breaks |  | 3.18 (0.89) | 2.87 (0.96) | 3.29 (0.94) |

BMI: body mass index; NSAID: non-steroidal anti-inflammatory drug; eGFR: estimated glomerular filtration rate; Tc: core body temperature.
\* Self-reported medication used in the last 24 hours. NSAID types included ibuprofen, aspirin, diclofenac, naproxen, or local brands that were identified as an NSAID.

Confounders assessed in this study included pre-shift eGFR, age, pre-shift serum osmolality, pre-shift creatine kinase, NSAID use, the company-reported amount of cane cut per day, work shift length, self-reported number of rest breaks during the workday, ambient HI (for the core-temperature models), ambient relative humidity, and the number of days between baseline and the study visit. Pre-shift osmolality and eGFR are measured before that day’s heat exposure and are treated as baseline covariates rather than same-shift mediators. PM is a plausible mediator of ambient heat and is therefore not adjusted for in the heat models; it is examined separately in the secondary bivariate analysis for the given observation. These confounders were used as adjustment factors in all models. Comorbid disease status including hypertension and diabetes was collected but not included as confounders because none of the participants met criteria for these conditions.

We estimated a marginal exposure-response curve (ERC) for each of the three heat metrics with a doubly-robust estimator,^50–51^ adjusting for confounders. The estimator projects a Neyman-orthogonal pseudo-outcome onto a low-dimensional natural-spline basis in the exposure. The pseudo-outcome combines an outcome-regression term with a generalized-propensity-score (density) reweighting term, so the curve is consistent when either nuisance model is correctly specified (double robustness). By default, the outcome model is an ordinary least squares spline in the exposure with linear covariates and the generalized propensity score is linear-normal; a more flexible machine-learning option is available. Uncertainty is a cluster-robust influence-function variance, clustered on the worker to account for repeated measures, combined across imputations by Rubin’s rules. Tabulated intervals are pointwise Wald limits, and the figure also shows a simultaneous 95% band over the exposure grid from a wild-cluster multiplier bootstrap. Time from baseline enters only as an adjustment covariate. Full details are in the Supporting Information.

We used a doubly-robust estimator to evaluate modified exposure policies (MEPs): hypothetical stochastic interventions that shift a worker’s exposure downward, and the expected change in cross-shift eGFR they would produce^52–53^. Identification requires conditional exchangeability (no unmeasured confounding given the observed covariates), consistency, and positivity. Because an MEP shifts each worker’s observed exposure rather than setting it to a fixed level, the shifted exposure stays within the support of the observed distribution, so positivity holds more readily than under static interventions. The DR estimator combines an outcome-regression model with density-ratio weights that reweight the observed exposure distribution to the shifted one, each estimated with a Super Learner ensemble (an intercept-only learner, elastic net, and multivariate adaptive regression splines); repeated measures are handled in the cluster-robust variance rather than the cross-validation folds. The density ratio is estimated by a classification approach: the observed data are duplicated with shifted exposure values, a classifier is fit to separate original from shifted observations, and its predicted odds recover the ratio.^51–52,54^ We tested additive shifts in 1-degree increments and multiplicative shifts in 1% increments for each heat metric. Each yields the average counterfactual change in eGFR under that reduction, contrasted against the null policy (the average observed outcome) to assess the plausibility and potential benefit of an intervention.

To manage missing exposure, outcome, and confounder data, multiple imputation by chained equations generated 50 imputed datasets. The imputation model included the outcome (pre- and post-shift eGFR), all three heat metrics, particulate matter, and every confounder used in the analysis models. Estimates from these datasets were combined according to Rubin’s rules^54^. Within-imputation variances use the cluster-robust influence-function estimator for both the exposure-response curve and the modified-exposure-policy analyses^52^.

A secondary bivariate MEP analysis assessed the individual and joint effects of PM and HI on cross-shift eGFR change.

All analysis was performed in R version 4.5.0. Additional details on the model construction are available in the *Supplemental Methods* section in the **Supporting Information, additional methods and results.**

## 3. Results

The study population (Table 1) consisted of 148 male workers with an average age of 31 years (SD: 10). At the time of enrollment, the cohort had an average body mass index of 23 kg/m^2^ (SD: 2.3), and an average eGFR of 115 mL/min/1.73m² (SD: 19.3). Throughout the harvest, increasing HI was observed, reaching an average of 38.2 °C (SD:1.09) at the end of the harvest. In contrast, the average daily Tc remained constant around 37.3 °C and the average change in Tc declined throughout the season.

Average pre-shift eGFR remained relatively constant across both seasons, and the cross-shift eGFR change was slightly smaller over the season.

As shown in Figure 1 and Table 2, the DR analysis showed exposure-dependent effects of heat on acute kidney function. Panel A shows the marginal exposure-response curve (ERC) for ambient HI, pooled over the harvest. The decline in eGFR steepened as HI increased. At an ambient HI of 34 °C, the estimated cross-shift change was −2.45 mL/min/1.73 m² (95% CI: −5.95, 1.05), which was not statistically significant, while at 40 °C the decline was statistically significant at −10.27 mL/min/1.73 m² (95% CI: −14.52, −6.02).

**Figure 1.**
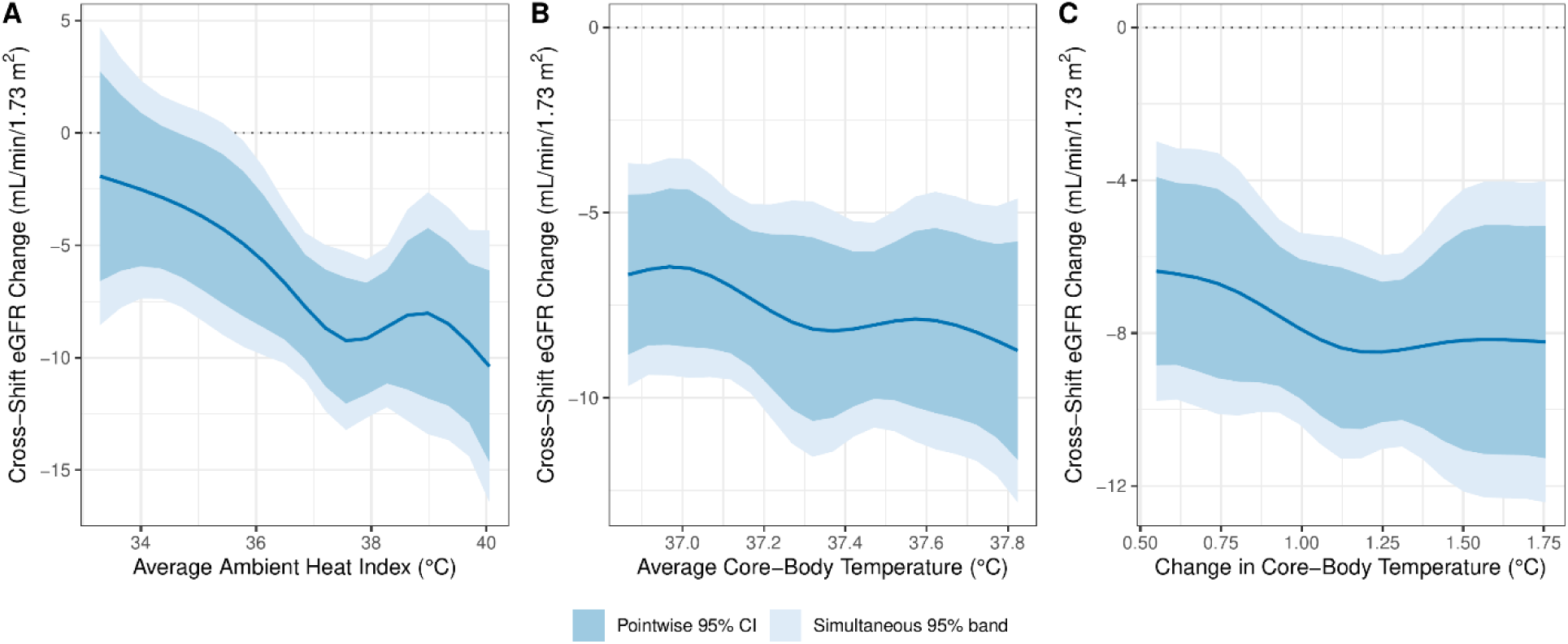
Marginal exposure-response curves (pooled over the harvest) for the change in eGFR from pre-shift to post-shift across levels of ambient HI, average daily Tc, and change in Tc (left to right), from the 10^th^ to the 90^th^ percentile, estimated with the doubly-robust estimator. Solid lines are the estimated curves. The darker shaded band is the pointwise 95% confidence interval and the lighter shaded band is the simultaneous 95% band from a wild-cluster multiplier bootstrap over the exposure grid. The dotted line marks no change.

**Table 2.**
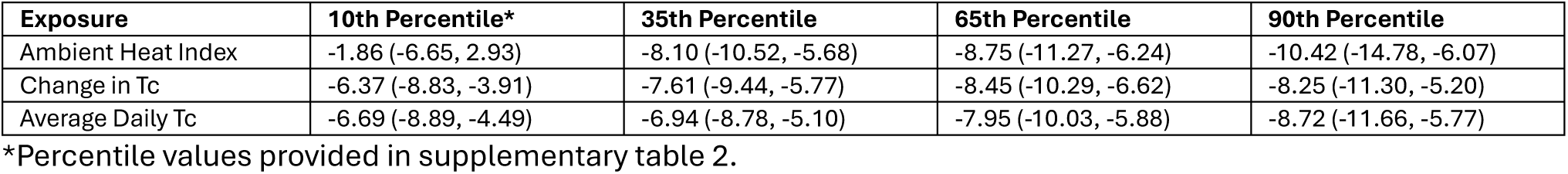
Doubly-robust estimates of the marginal exposure-response curves (pooled over the harvest), showing the effect of Heat Index (HI), average daily core body temperature (Tc), and change in Tc on cross-shift eGFR change at the 10^th^, 35^th^, 65^th^, and 90^th^ percentiles of each exposure, with pointwise 95% confidence intervals.

Panels B and C show the marginal ERCs for average Tc and change in Tc. Both were negative across their observed ranges, indicating that higher core temperature was associated with greater cross-shift eGFR decline. At the 65th percentile of average Tc the estimated change was −7.95 mL/min/1.73 m² (95% CI: −10.03, −5.88). Average workday Tc exceeded 37.2 °C in 57% of worker-days.

Figure 2 panels A, B, and C show the results of additive shifts in heat exposure and panels D, E, and F show the results of multiplicative shifts in heat exposure. Reducing HI exposure by 2 °C (panel A) would reduce eGFR decline by 3.00 mL/min/1.73 m² (95% CI: 0.92, 5.09). Similarly, reducing HI exposure by 5% (panel D) reduced eGFR decline by 2.94 mL/min/1.73 m² (95% CI: 0.96, 4.91). Additive and multiplicative reductions in Tc did not produce statistically significant changes in eGFR, despite the negative Tc curves, because the feasible shifts move core temperature only slightly. Sensitivity analyses using raw serum creatinine and cystatin-C-based eGFR, reported in the Supporting Information, were directionally consistent with these results.

**Figure 2.**
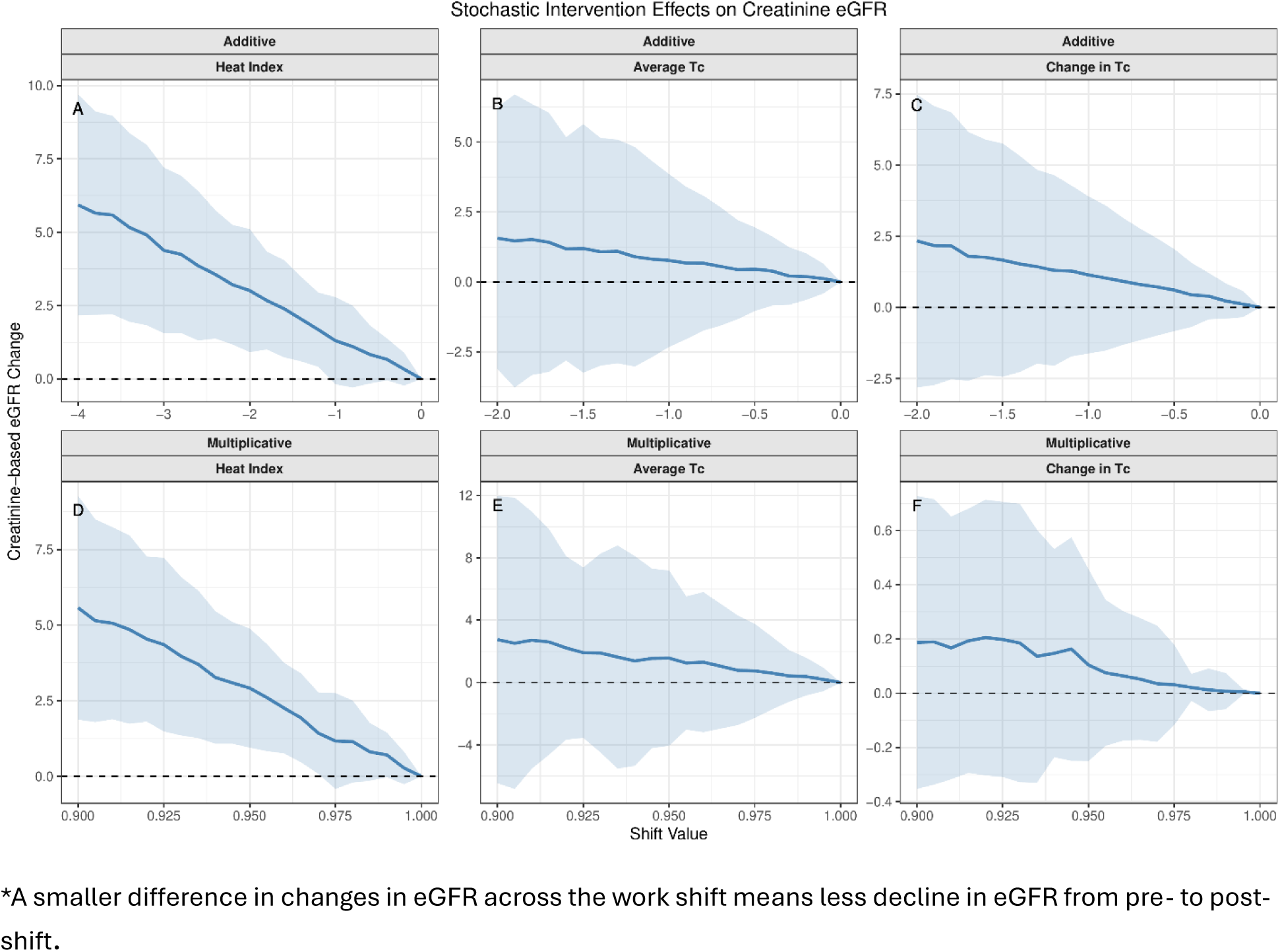
The figure displays the differences in changes in eGFR across the work shift when average ambient HI, change in Tc, and average daily Tc (left to right) were restricted to the heat metric levels along the x-axis as part of the modified intervention policy change. The dashed line at zero represents no expected change in eGFR across the work shift. *A smaller difference in changes in eGFR across the work shift means less decline in eGFR from pre- to post-shift.

The results of the secondary analysis revealed that HI alone had a negative impact on kidney function, as shown in the primary analysis. In contrast, there was no significant impact of PM either individually or jointly on the outcome. A 10% multiplicative shift of PM exposure produced a near-zero, non-significant change of −0.05 mL/min/1.73 m² (95% CI: −0.41, 0.32). Similarly, a ½ standard deviation additive shift produced a non-significant change of 0.11 mL/min/1.73 m² (95% CI: −0.82, 1.05). Both estimates are statistically indistinguishable from zero. Details on these results can be found in the supporting Information, additional methods and results.

## 4. Discussion

This study provides causal evidence that agricultural workers can experience clinically meaningful declines in kidney function while performing physically demanding work under extreme heat. These declines, roughly 2 to 10 mL/min/1.73 m² of eGFR across the range of heat index, are on the order of the cross-shift changes linked to incident kidney injury in this workforce^11^, and they accrue across a six-month harvest. Using doubly-robust methods, higher ambient HI was associated with worsening kidney function across a work shift among male Guatemalan sugarcane harvesters, consistent with a causal effect. Higher core temperature was also associated with acute declines in kidney function.

In this cohort, higher ambient HI was associated with worse kidney function across the work shift. This is consistent with proposed mechanisms in which repeated heat exposure and volume depletion lead to tubular injury, with vasopressin and uric-acid pathways implicated^55^. Heat acclimatization is thought to attenuate heat-induced kidney injury and additional studies are needed to evaluate the timing, mechanisms, and effectiveness of current acclimatization practices^56^. Acclimatization may involve physiological adaptations such as better fluid and electrolyte balance^57^ and improved cardiovascular function^58^. These mechanisms could reduce both fluid imbalances and cardiovascular stress, thereby lowering the risk of kidney injury in workers engaged in heavy manual labor such as sugarcane harvesting.

Through the MEP analysis, we identified policy-relevant reductions in ambient heat exposure expected to improve worker health. Specifically, limiting ambient heat index by 5% was estimated to reduce cross-shift declines in kidney function. Such an intervention could take different forms such as shifting work hours or more closely monitoring ambient heat to slow or stop work under hazardous conditions. Future studies, guided by implementation science frameworks, are needed to design and evaluate the feasibility, uptake, and effectiveness of such interventions in occupational settings. Evaluating the short and long-term clinical relevance of this potential reduction in daily decrements to kidney function is warranted.

Higher average workday Tc was associated with modest cross-shift declines in eGFR across the harvest. In these workers, average workday Tc was frequently above the “normal” range (36.1 to 37.2 °C). Factors such as high ambient temperatures, elevated humidity, and dehydration can compromise thermoregulation and hinder the body’s capacity to maintain a stable internal temperature. Future research should further investigate these associations and the mechanisms linking impaired thermoregulation and kidney function decline.

While the average Tc is an intuitive summary statistic, Tc naturally fluctuates throughout the day. Large Tc fluctuations have been associated with negative health impacts such as increased cardiovascular stress and impaired central nervous system functioning^19^.

Therefore, the change in Tc was used to capture the amount of Tc fluctuation during the workday, relative to the worker’s baseline Tc. The results suggest that greater cross-shift change in Tc was causally related to eGFR decline. Previous studies have established that heat strain, characterized by a combination of Tc and heart rate, has been linked to worsening kidney function in agricultural communities; because we measured Tc but not heart rate, we capture only one component of that definition^59^. This study’s findings align with this evidence and suggest that internal heat strain may be a driver of kidney damage, and that interventions specifically addressing heat strain are needed to reduce the risk of CKDu^4,23,60^.

The present data suggest that high heat exposure can lead to kidney function decline and that reducing outdoor HI exposure by 5% may reduce acute injury to the kidney during work shifts. Previous research has shown that working in temperatures greater than 35°C increases risk of heat stress^17^. It has been hypothesized that repeated heat stress and recurrent bouts of acute kidney injury contribute to CKDu^61–62^, although direct support of causation is limited. These findings support implementing heat standards that limit exposure and protect workers across the harvest^63^.

This study’s findings reinforce that heat is a workplace hazard, corroborating prior studies^64 65^. However, few workplace intervention studies have evaluated strategies to address this hazard^38–39,59,66–70^. The results of this study provide evidence to guide the design and timing of workplace interventions to mitigate the harmful effects of heat. These findings suggest that reducing ambient heat exposure can lower acute kidney injury risk across the harvest. The core-temperature curves point in the same direction, but feasible reductions in core temperature did not produce statistically significant benefits, which raises questions about how best to lower internal heat strain.

In a secondary analysis, we observed no statistically significant effect of PM exposure on kidney function, either independently or jointly with HI. While these results do not definitively exclude PM or PM components such as silica and metals, they do reinforce the need to focus effort on high heat exposure to protect kidney health. While heat is an acute stressor, PM exposure has been linked to chronic inflammation over time^71–72^. Since this study looked at acute changes in kidney function, latent or long-term consequences of PM may not have been detected. The PM5 fraction bears on this as well. It includes the fine particles most often tied to systemic and renal effects, but on a mass basis it is weighted toward coarser particles that deposit in the upper airway and are less likely to reach the kidney. With the high first-visit missingness and the modest sample, the secondary PM analysis is likely underpowered, so the null should not be read as evidence that PM has no effect. Future studies should look at the effects of PM on long-term kidney outcomes. In addition, high PM exposures experienced by these workers suggest the need for research on other health endpoints including cardiovascular, respiratory, cognitive, and malignant disorders. Taken together, the results of the primary and secondary analysis suggest two future directions of occupational health and safety in this population. First, the implementation of various interventions are needed to protect workers from extreme heat. Second, more research is needed to characterize and investigate the health impacts of other environmental hazards including PM and PM components.

### 4.1. Causal Inference Approach to Policy Development

The present study is among the first to apply modified exposure policies to occupational heat exposure and kidney function, building on prior applications of g-methods in environmental and occupational health. The two estimands answer different questions. The exposure-response curve describes how kidney function varies with the level of an exposure, which is useful for understanding the hazard and for setting exposure limits, yet it corresponds to fixing every worker at a common level, a hypothetical intervention that need not be achievable in practice. A modified exposure policy instead shifts each worker’s own exposure by a realistic amount, so the counterfactual stays within the observed range and the estimand corresponds to an intervention a worksite could adopt, such as reducing HI by 5% during work shifts. For this reason, the policy analysis provides more direct evidence for the potential benefit of workplace heat standards and related public health guidelines (see the Supporting Information, additional methods and results, for further details).

Given CKDu’s complex etiology, the same policy approach extends to other suspected risk factors. Future studies could evaluate interventions that reduce air pollutant and heavy-metal exposures, both thought to contribute to CKDu in agricultural communities^73–74^, and could compare the benefit of targeting different exposures to help prioritize among them. Because a modified exposure policy is defined as a realistic shift in exposure, it names a specific intervention that can be carried into a field trial or natural experiment. We note, however, that the health improvements estimated under these modeled policies are hypothetical and at times rest on stringent assumptions. We therefore do not present the approach as a replacement for the randomized trials and natural experiments that remain the standard for confirming whether an intervention delivers the expected benefit; it identifies and quantifies the interventions most worth testing.

### 4.2. Limitations

These methods strengthen, but do not by themselves guarantee, a causal interpretation, which remains subject to unmeasured confounding, selection, and measurement error. This study was conducted at a worksite with an established health and safety culture, including longstanding programs to address heat stress and kidney disease and a hiring screen that excludes applicants with poor kidney function. The cohort is therefore healthier than the general harvester population, which would tend to make the observed declines underestimate the true effect, and the results may not generalize to other worker populations. Greater loss of eGFR has been observed in previous studies when workers with more marginal kidney function were permitted to work^67^, prompting speculation that a less healthy workforce may experience an even greater loss of kidney function under similar conditions of heat and PM exposure. It is concerning, and noteworthy, that we are observing impacts on kidney health in a largely healthy cohort. The exposure is summarized as a single daily value, so the policy analysis cannot represent the timing or duration of peak exposure within a shift. For example, a worker exposed to high temperatures for a few minutes may experience less kidney function decline than a worker exposed to high temperatures for a few hours. More work is needed to test the effectiveness of interventions targeting heat exposure.

This study uses serum creatinine to calculate eGFR, which is then used as a measure of kidney function. The authors recognize that changes in serum creatinine are a surrogate for kidney tissue injury. Heat index omits the radiant and air-movement terms of WBGT and serves as a proxy for occupational heat stress rather than the occupational standard. Additionally, the measurement of PM has limitations including minor issues with data collection, unanticipated variability between timepoints due to environmental events such as rain, and modest differences between pre-shift and post-shift measurements^28^. These limitations, however, are small relative to the overall magnitude of exposure and do not fully explain the observed biological responses. Instead, the challenge lies in distinguishing the PM-associated inflammatory effects, which may be best evaluated through mediation analyses or metabolomics.

While we attempt to model these relationships without bias, unmeasured confounding may have biased the results. Researchers should carefully consider other co-exposures and unmeasured confounders when extending this approach to analyzing exposure-outcome relationships, including other environmental factors such as silica and other nephrotoxins. In addition, this study had a limited sample size (148 workers) and substantial missing data, especially for the core-temperature metrics (about 50% per visit) and first-visit particulate matter (about 64%); these are quantified in the Supporting Information. As a secondary analysis of an existing cohort, no a priori power calculation was performed, and the wide confidence intervals reflect the available precision. Attrition across the harvest (from 148 to 115 workers) may be informative, which multiple imputation cannot fully address. Cross-shift creatinine can also rise from hemoconcentration as well as from reduced filtration; we adjust for morning osmolality, but this may not fully separate the two, and a cystatin-C-based sensitivity analysis, reported in the Supporting Information, was directionally consistent with the creatinine-based results but less precise. Despite these challenges, incorporating stochastic along with fixed evaluations of an exposure-response into environmental health studies offers a practical option for translating epidemiologic findings into actionable strategies. This study’s findings illustrate this approach by demonstrating how reductions in heat exposure may prevent kidney injury and mitigate other environmentally driven health risks.

## 5. Conclusion

This study provides evidence that increases in ambient HI and Tc, evaluated separately, are associated with declines in kidney function. It shows that workplace interventions to reduce heat exposure may lower the risk of kidney injury. By pairing the exposure-response curve with modified exposure policies, the analysis both characterizes the heat-kidney relationship and quantifies the potential benefit of realistic reductions in exposure. These doubly-robust methods offer a transparent basis for designing and evaluating heat-reducing interventions as extreme heat becomes more frequent, while recognizing that they cannot by themselves establish causation.

## Data Availability

Data and statistical code used in this study is available upon request.

## Acknowledgement

We would like to acknowledge our Guatemalan collaborators, particularly the participating workers who made this research possible.

