## Supplement for "Policy-Relevant Causal Approach to Assessing the Impact of Occupational Heat and Airborne Particulate Matter Exposure on Acute Kidney Function in Guatemalan Sugarcane Workers"

### Supplemental Methods

*Doubly-Robust Exposure-Response Curve (ERC) Estimator*

We estimated the marginal exposure-response curve (ERC) with a doubly-robust estimator, replacing the plug-in generalized additive model g-computation used previously. Let $A$ denote a heat metric (ambient heat index, average core temperature, or change in core temperature), $Y$ the cross-shift change in eGFR, and $\mathbf{W}$ the vector of baseline and time-varying covariates. Writing $Y(a)$ for the potential outcome under exposure level $a$, the estimand is the average potential outcome,

$$\theta(a) = \mathrm{E}\{Y(a)\}.$$

Identification of $\theta(a)$ requires consistency, conditional exchangeability given $\mathbf{W}$ (no unmeasured confounding), and positivity.

Let $\mu(a, \mathbf{w}) = \mathrm{E}(Y \mid A = a, \mathbf{W} = \mathbf{w})$ be the outcome regression and $g(a \mid \mathbf{w})$ the conditional density of the exposure given covariates (the generalized propensity score). Define the covariate-marginalized curve $\bar{\mu}(a) = \mathrm{E}_{\mathbf{W}}\{\mu(a, \mathbf{W})\}$, which is the plug-in g-computation curve. Following the continuous-treatment construction of Kennedy and colleagues[8], the doubly-robust pseudo-outcome is

$$\xi_i = \bar{\mu}(A_i) + \frac{\varpi(A_i)}{g(A_i \mid \mathbf{W}_i)}\{Y_i - \mu(A_i, \mathbf{W}_i)\},$$

where $\varpi(a)$ is the marginal exposure density that stabilizes the weights. The first term is the plug-in g-computation; the second is the orthogonalizing inverse-probability residual. Because the pseudo-outcome is Neyman-orthogonal, $\hat{\theta}(a)$ is consistent if either $\mu$ or $g$ is correctly specified (double robustness).

We projected the pseudo-outcome onto a fixed natural-spline working model $b(a) = \left(1, b_1(a), \dots, b_K(a)\right)^T$, in the spirit of the debiased series projection of Semenova and Chernozhukov[9]. The working model is $\theta(a) = b^T(a)\beta$, estimated by ordinary least squares on the pseudo-outcomes,

$$\hat{\beta} = \left\{\sum_{i=1}^{n} b(A_i)\, b^T(A_i)\right\}^{-1} \sum_{i=1}^{n} b(A_i)\,\xi_i, \qquad \hat{\theta}(a) = b^T(a)\hat{\beta}.$$

By default the nuisance functions use a parsimonious parametric specification: $\mu$ is an ordinary least squares regression that is a spline in the exposure plus linear covariate terms, and $g$ is a linear-normal, homoscedastic model. A more flexible machine-learning option (a Super Learner ensemble) is available. The parametric default is preferred here because flexible density estimates can become very small, inflating the stabilized weights $\varpi/g$ and the variance at this sample size.

**Variance.** Inference is cluster-robust at the worker level. The influence-function contribution for observation $i$ is

$$\psi_i = b(A_i)\left\{\xi_i - b^T(A_i)\hat{\beta}\right\} + \zeta_i, \qquad \zeta_j = \frac{1}{n}\sum_{i=1}^{n} b(A_i)\ \left\{\mu\left(A_i, \mathbf{W}_j\right) - \bar{\mu}(A_i)\right\}.$$

The $\zeta_j$ term is the additional influence from marginalizing $\mu$ over the covariate distribution; omitting it understates the variance. Let workers (clusters) be indexed by $c$ with $U_c = \sum_{i\in c} \psi_i$, and let $M_n = \sum_i b\,(A_i)\ b^T(A_i)$. The cluster-robust (CR1) covariance is

$$\widehat{\mathrm{Var}}(\hat{\beta}) = M_n^{-1}\left(\sum_c U_c\ U_c^T\right)M_n^{-1}, \qquad \widehat{\mathrm{Var}}\left\{\hat{\theta}(a)\right\} = b^T(a)\widehat{\mathrm{Var}}\left(\hat{\beta}\right) b(a).$$

Pointwise 95% intervals use Wald limits; simultaneous bands over the exposure grid use a wild-cluster multiplier bootstrap. Time from baseline enters only as an adjustment covariate, so the reported curve is marginal and pooled over the harvest. Estimates and variances are combined across the 50 multiply imputed datasets by Rubin's rules.

*Modified Exposure Policy Analysis via Doubly Robust (DR) Estimator*

Alongside the doubly-robust exposure-response curve described above, we evaluated modified exposure policies (MEPs) with a separate doubly-robust (DR) estimator. An MEP is a hypothetical intervention that modifies the exposure in a way that can depend on its natural (observed) value. In our context, we defined MEPs as additive and multiplicative shift interventions on temperature.

The causal quantity of interest for each policy was the mean change in eGFR under that intervention, denoted $\psi_\delta = E\left[Y\left(A^\delta\right)\right]$ where $Y\left(A^\delta\right)$ denotes the counterfactual outcome if the exposure were modified according to policy $\delta$. We contrasted an estimate of this quantity with the no-intervention (observed exposure) scenario. Let $\psi_0 = E[Y] = E[Y(A)]$ be the population mean outcome under the observed exposure distribution (this is estimated by the sample mean change in eGFR). We define the causal effect of the policy as the difference in mean outcome compared to no policy implementation: $\tau_\delta = \psi_\delta - \psi_0$. This contrast $\tau_\delta$ is a population average stochastic intervention, measuring how the mean outcome would change if the exposure were shifted by $\delta$ versus the status quo.

To estimate $\psi_\delta$, each worker-day was treated as a single observation, containing an exposure $A$ (ambient or core-body temperature) and outcome $Y$ (same-day eGFR change), with baseline and daily (time-varying) covariates $\mathbf{W}$ to adjust for confounding. We assume that conditional on the observed covariates, there is no unmeasured confounding for the exposure-outcome relationship (exchangeability), and we also assume consistency and positivity. Under these assumptions, $\psi_\delta$ is identified by the g-formula for the MEP.[3,4]

*Density Ratio Estimation.* A central component of our doubly robust estimator is the density ratio, defined as

$$r_\delta(a, \mathbf{w}) = \frac{g^\delta(\, a \mid \mathbf{w}\,)}{g(\, a \mid \mathbf{w}\,)},$$

where $g(a \mid \mathbf{w})$ denotes the conditional density of the observed exposure given covariates, and $g^\delta(a \mid \mathbf{w})$ denotes the density of the exposure under the shifted policy $\delta$.[3,7] Rather than estimating the numerator and denominator densities separately, we employed a classification-based approach to estimate the density ratio directly.[7] This approach proceeds by constructing an augmented dataset of size $2N$: the original observations with their observed exposures $A_{ij}$ and a duplicated set of observations where the exposure is replaced by its shifted value $A_{ij}^\delta$. A binary label $D$ is assigned to each observation, where $D = 0$ for original observations and $D = 1$ for duplicated observations. Within this augmented dataset, the covariates $\mathbf{W}_{ij}$ are identical across original and duplicated copies for the same individual-time pair.

A classification model is then fit to predict $\Lambda$ from the exposure and covariates $(A, \mathbf{W})$ in this augmented dataset. The estimated density ratio is recovered from the predicted conditional odds:

$$\hat{r}_\delta(a, \mathbf{w}) = \frac{\hat{P}(\, D = 1 \mid a, \mathbf{w}\,)}{\hat{P}(\, D = 0 \mid a, \mathbf{w}\,)}.$$

This classification approach avoids the challenge of directly estimating continuous conditional densities and allows the use of flexible machine learning algorithms for the density ratio.[7] We estimated density ratios using the lmtp R package,[7] which implements this classification-based density ratio procedure internally. For the density ratio classifier, we specified a Super Learner ensemble library consisting of an intercept-only (mean) regression, regularized regression (elastic net via glmnet), and multivariate adaptive regression splines (MARS via the earth package). Each learner within the Super Learner ensemble uses 10-fold cross-validation internally for ensemble weight selection.

*Outcome Model.* Separately, we estimated the outcome regression $\mu(A, \mathbf{W}) = E[Y \mid A, \mathbf{W}]$ using a Super Learner ensemble with the same library of learners: an intercept-only regression, elastic net, and MARS.

*Doubly Robust Estimator.* We combined the density ratio estimate and the outcome model within a doubly robust estimating framework. For each shift policy $\delta$, the efficient influence function (EIF) for the single time-point setting takes the form[3,4,7]

$$\varphi_\delta(A, \boldsymbol{W}, Y) = r_\delta(A, \mathbf{W})\{Y - \mu(A, \mathbf{W})\} + \mu\left(A^\delta, \mathbf{W}\right) - \psi_\delta.$$

This estimating equation motivates the doubly robust point estimate

$$\hat{\psi}_\delta = \frac{1}{N} \sum_{i,j} \left[\hat{r}_\delta\left(A_{ij}, \mathbf{W}_{ij}\right)\left\{Y_{ij} - \hat{\mu}\left(A_{ij}, \mathbf{W}_{ij}\right)\right\} + \hat{\mu}\left(A_{ij}^\delta, \mathbf{W}_{ij}\right)\right],$$

where $\hat{r}_\delta(A_{ij}, \mathbf{W}_{ij})$ is the estimated density ratio and $\hat{\mu}$ is the fitted outcome regression. The estimate $\hat{\tau}_\delta = \hat{\psi}_\delta - \bar{Y}$ is then the estimated causal contrast, with $\bar{Y} = N^{-1} \sum_{i,j} Y_{ij}$ being the sample mean outcome. This two-stage procedure, wherein the density ratios are extracted from lmtp and the outcome model is fit separately via Super Learner, ensures that each nuisance function is estimated with a tailored model while retaining the doubly robust property of the combined estimator.

The estimator is doubly robust in the sense that it remains consistent if either the density ratio or the outcome model is consistently estimated, providing protection against model misspecification.[3,4] Because both nuisance functions are estimated with flexible Super Learner ensembles, the estimator also benefits from rate double robustness: the estimation error is a product of the errors in the two nuisance functions, which can be small even when both models are imperfectly estimated.[7]

*Variance Estimation.* We used a cluster-robust sandwich estimator based on the influence function. Define the estimated influence function values

$$\hat{\varphi}_{ij} = \hat{r}_\delta(A_{ij}, \mathbf{W}_{ij})\{Y_{ij} - \hat{\mu}(A_{ij}, \mathbf{W}_{ij})\} + \hat{\mu}(A_{ij}^\delta, \mathbf{W}_{ij}) - \hat{\psi}_\delta.$$

The cluster-robust variance of $\hat{\tau}_\delta$ is then

$$\widehat{Var}(\hat{\tau}_\delta) = \frac{n}{(n-1)N^2} \sum_{i=1}^{n} \left( \sum_{j=1}^{m_i} \hat{\varphi}_{ij} - Y_{ij} + \bar{Y} \right)^2,$$

where $n$ denotes the number of independent clusters (workers) and $N$ denotes the total number of observations. Confidence intervals were constructed at the 95% level using Wald-type intervals.

*Bivariate Modified Exposure Policy Analysis*

We extended the univariate doubly robust MEP framework to accommodate bivariate exposures, enabling estimation of the individual and joint effects of two concurrent environmental exposures on kidney function. In our secondary analysis, the two exposures of interest were particulate matter (PM; denoted $A_1$) and heat index (HI; denoted $A_2$), with the outcome $Y$ being the cross-shift change in eGFR.

*Estimands.* Let $\delta_1$ and $\delta_2$ denote the shift magnitudes for PM and HI, respectively. Under additive shift policies, the shifted exposures are $A_1^{\delta_1} = A_1 - \delta_1$ and $A_2^{\delta_2} = A_2 - \delta_2$; under multiplicative shift policies, $A_1^{\delta_1} = A_1 \times \delta_1$ and $A_2^{\delta} = A_2 \times \delta_2$. We defined four estimands to characterize the bivariate exposure-response relationship in terms of the additive shifts:

1. *Joint effect:* The expected change in eGFR when both exposures are simultaneously shifted, contrasted against the observed mean:

$$\tau_{\text{joint}} = E\left[Y\left(A_1^{\delta_1}, A_2^{\delta}\right) - Y\right] = \psi_{\text{joint}} - E[Y].$$

2. *Marginal effect of $A_1$ (PM):* The expected change in eGFR when only PM is shifted, holding HI at its observed level:

$$\tau_{\delta_1} = E\left[Y\left(A_1^{\delta_1}, \mathrm{A}_2\right) - Y\right] = \psi_{\delta_1} - E[Y].$$

3. *Marginal effect of $A_2$ (HI):* The expected change in eGFR when only HI is shifted, holding PM at its observed level:

$$\tau_{\delta_2} = E\left[Y\left(A_1, A_2^{\delta}\right) - Y\right] = \psi_{\delta_2} - E[Y].$$

4. *Interaction:* The extent to which the joint effect departs from the sum of the two marginal effects:

$$\tau_{\text{int}} = \tau_{\text{joint}} - \tau_{\delta_1} - \tau_{\delta_2}.$$

A positive interaction indicates synergism (the joint reduction in exposure produces a larger health benefit than the sum of the individual reductions), while a negative interaction indicates antagonism.

*Estimation.* The joint effect was estimated using the bivariate extension of the density ratio-based doubly robust procedure described above. Specifically, we estimated a density ratio for the bivariate shifted exposure $\left(A_1^{\delta_1}, A_2^{\delta_2}\right)$ via the lmtp package, treating both exposures jointly as the treatment vector, and fit an outcome model $\hat{\mu}(A_1, A_2, \mathbf{W})$ via Super Learner with both exposures as predictors. The joint doubly robust estimate takes the form

$$\hat{\psi}_{\text{joint}} = \frac{1}{N}\sum_{i,j}\left[\hat{r}_{\delta_1,\delta_2}(A_{1,ij}, A_{2,ij}, \mathbf{W}_{ij})\{Y_{ij} - \hat{\mu}(A_{1,ij}, A_{2,ij}, \mathbf{W}_{ij})\} + \hat{\mu}\left(A_{1,ij}^{\delta_1}, A_{2,ij}^{\delta_2}, \mathbf{W}_{ij}\right)\right].$$

Similar estimates for the marginal exposure effects $\hat{\psi}_{\delta_1}$ and $\hat{\psi}_{\delta_2}$ are obtained by applying the appropriate shift only to the respective exposure, $A_1^{\delta_1}$ or $A_2^{\delta_2}$, and applying the correct density ratio, $\hat{r}_{\delta_1}$ or $\hat{r}_{\delta_2}$. These estimates can be used to find $\hat{\tau}_{\text{joint}} = \hat{\psi}_{\text{joint}} - \bar{Y}$, $\hat{\tau}_{\delta_1} = \hat{\psi}_{\delta_1} - \bar{\mathrm{Y}}$, $\hat{\tau}_{\delta_2} = \hat{\psi}_{\delta_2} - \bar{\mathrm{Y}}$, and $\hat{\tau}_{\text{int}} = \hat{\psi}_{\text{joint}} - \hat{\psi}_{\delta_1} - \hat{\psi}_{\delta_2} + \bar{\mathrm{Y}}$

For the additive shift analysis, shift magnitudes were set to $\delta_1 = 0.5 \times \mathrm{SD}(A_1)$ and $\delta_2 = 0.5 \times \mathrm{SD}(A_2)$, representing a half-standard-deviation reduction in each exposure. For the multiplicative shift analysis, both exposures were reduced by a fixed percentage (e.g., $\delta = 0.95$ for a 10% reduction). These shift magnitudes were selected to represent practically meaningful reductions while remaining within the support of the observed exposure distributions to satisfy the positivity assumption.[3,4]

*Multiple Imputation.* To manage missing exposure, outcome, and confounder data across both the univariate and bivariate analyses, multiple imputation by chained equations was implemented to generate 50 imputed datasets. Estimates from these datasets were then combined according to Rubin's rules.[6] Within-imputation standard errors were calculated using the cluster-robust influence function-based variance estimator described above. Between-imputation variance was estimated as the sample variance of the point estimates across the 50 imputations. The total variance combined both components using the standard formula $T = \bar{U} + (1 + 1/m)B$, where $\bar{U}$ is the average within-imputation variance, $B$ is the between-imputation variance, and $m$ is the number of imputations.

**Heat index calculation.** Heat index was computed from air temperature $T$ (in °F) and relative humidity $R$ (in %) using the National Weather Service Rothfusz regression,

$$\mathrm{HI} = -42.379 + 2.04901523\,T + 10.14333127\,R - 0.22475541\,TR$$

$$-0.00683783\,T^2 - 0.05481717\,R^2 + 0.00122874\,T^2R$$

$$+0.00085282\,TR^2 - 0.00000199\,T^2R^2,$$

with the standard adjustments at low relative humidity and at low temperature. Heat index was then converted to °C.

### Companion Analysis: Raw Serum Creatinine

Serum creatinine underlies the estimated glomerular filtration rate (eGFR), yet eGFR assumes a steady state that a single work shift may not meet. As a companion to the primary analysis, we estimated the exposure-response curves for the cross-shift change in raw serum creatinine measured on the iSTAT point-of-care device. The estimator, confounder set, and Winsorization match the eGFR analysis, and the outcome is imputed in a model separate from the creatinine-derived eGFR. Creatinine rose with heat across all three metrics, mirroring the eGFR decline: each curve is positive and steepens over the observed exposure range (Table S4).

We repeated the modified exposure policy (MEP) analysis with raw creatinine as the outcome (Figure S2, Table S5). A 2 °C or 5% reduction in Heat Index lowered the cross-shift rise in creatinine, consistent with the eGFR gains in the main text. Reductions in average or change in Tc moved creatinine in the same protective direction, yet neither reached significance, matching the eGFR shift results.

### Companion Analysis: Cystatin-C

Cystatin C is a filtration marker largely independent of muscle mass, so it provides a check on the creatinine-based results. We repeated the exposure-response and modified-exposure-policy analyses using cystatin-C-based eGFR and raw serum cystatin C, each imputed separately (Figure S1, Tables S4 and S5). The cystatin-C estimates were directionally consistent with the creatinine-based primary analysis, with kidney function declining across the heat range and improving under simulated reductions, but none reached statistical significance. This imprecision is consistent with the high cystatin-C missingness at the later visits (Table S1). The cystatin-C analyses therefore neither contradict nor independently confirm the primary results.

### Supplemental Figures and Tables

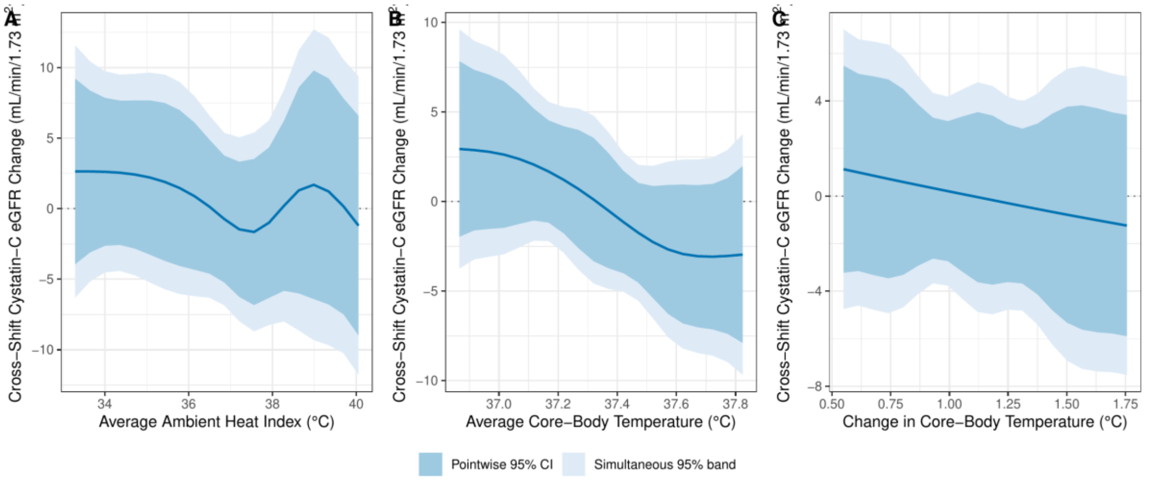


Figure S1. Doubly-robust marginal exposure-response curves for the cross-shift change in cystatin-C-based eGFR (mL/min/1.73 m2) across ambient heat index, average daily core-body temperature, and change in core-body temperature (left to right), from the 10th to the 90th percentile. The darker band is the pointwise 95% confidence interval and the lighter band is the simultaneous 95% band. The dotted line marks no change. This is the cystatin-C companion to Figure 1 and Table S4.

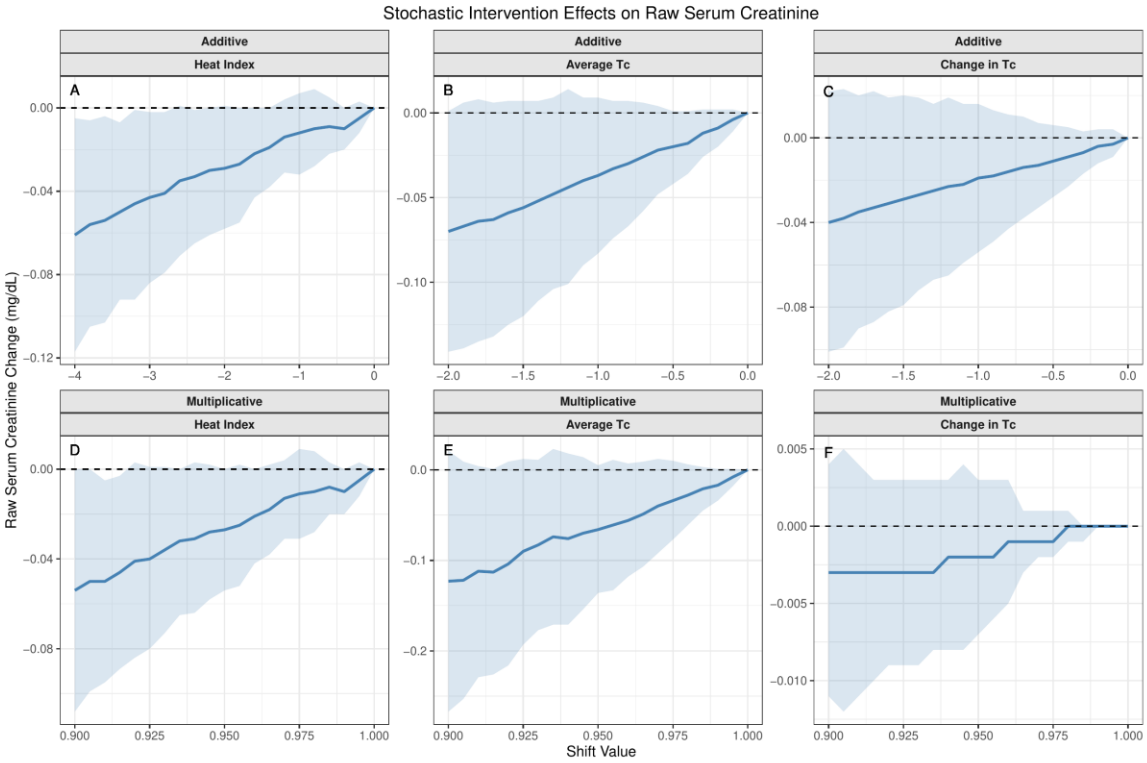


Figure S2. Doubly-robust modified exposure policy (MEP) estimates for the cross-shift change in raw serum creatinine (mg/dL) under additive (top row) and multiplicative (bottom row) reductions in ambient heat index, average daily core-body temperature, and change in core-body temperature. The dashed line marks no change, and negative values indicate a smaller rise in creatinine. This is the raw-creatinine companion to Figure 2 and Table S5.

**Table S1. Percent of worker-days missing by analysis variable and visit (Visit 1 N=145, Visit 2 N=118, Visit 3 N=115). Missingness is highest for the core-temperature metrics and for first-visit particulate matter.**

| Variable | Visit 1 | Visit 2 | Visit 3 |
|---|---|---|---|
| Cross-shift eGFR change (outcome) | 5% | 3% | 5% |
| Ambient heat index | 9% | 36% | 0% |
| Average Tc | 50% | 49% | 57% |
| Change in Tc | 50% | 50% | 57% |
| Particulate matter (PM5) | 64% | 2% | 3% |
| Morning osmolality | 2% | 0% | 1% |
| Morning creatine kinase | 2% | 0% | 3% |
| Company-reported cane cut | 45% | 9% | 10% |
| Shift length | 0% | 0% | 1% |
| Rest breaks | 0% | 0% | 2% |
| Relative humidity | 9% | 36% | 0% |
| NSAID use | 10% | 8% | 9% |
| Cystatin-C eGFR (auxiliary) | 1% | 64% | 2% |

Table S2. Percentile values for ambient HI, change in Tc, and average daily Tc. Percentiles are pooled over all three timepoints.

| Exposure | 10th Percentile | 35th Percentile | 65th Percentile | 90th Percentile |
|---|---|---|---|---|
| Ambient Heat Index | 33.3 | 37.0 | 38.2 | 40.1 |
| Change in Tc | 0.55 | 0.94 | 1.29 | 1.76 |
| Average Daily Tc | 36.9 | 37.1 | 37.5 | 37.8 |

Table S3. Results of secondary analysis, bivariate MEP relating particulate matter and heat index to cross-shift changes in eGFR.

| Shift Type | Parameter | Estimate | 95% CI Lower | 95% CI Upper |
|---|---|---|---|---|
| Multiplicative | Joint | 5.87 | 2.25 | 9.50 |
| Multiplicative | Particulate Matter | -0.05 | -0.41 | 0.32 |
| Multiplicative | Heat Index | 5.77 | 2.04 | 9.51 |
| Multiplicative | Interaction | 0.15 | -0.79 | 1.08 |
| Additive | Joint | 1.51 | -0.07 | 3.10 |
| Additive | Particulate Matter | 0.11 | -0.82 | 1.05 |
| Additive | Heat Index | 1.27 | -0.06 | 2.60 |
| Additive | Interaction | 0.13 | -0.67 | 0.92 |

*Note: Multiplicative shifts correspond to a 10% proportional reduction in each exposure (i.e., shifted exposure = observed exposure × 0.90). Additive shifts correspond to a half-standard-deviation absolute reduction in each exposure (i.e., shifted exposure = observed exposure − 0.5 × SD). Joint effects reflect simultaneous shifts in both particulate matter and heat index; marginal effects reflect shifting one exposure while holding the other at its observed level. The interaction term quantifies departure from additivity of the two marginal effects.*

Table S4. Doubly-robust exposure-response curves for the cross-shift change in four kidney-function outcomes at the 10th, 35th, 65th, and 90th percentiles of Heat Index, change in core body temperature (Tc), and average daily Tc, with 95% confidence intervals. The eGFR outcomes are in mL/min/1.73 m2, where a negative value is a decline; the raw markers are in mg/dL (creatinine) and mg/L (cystatin C), where a positive value is a rise. The creatinine-based eGFR rows reproduce Table 2 in the main text.

| **Outcome** | **Exposure** | **10th Percentile** | **35th Percentile** | **65th Percentile** | **90th Percentile** |
|---|---|---|---|---|---|
| Creatinine eGFR (mL/min/1.73 m2) | Ambient Heat Index | -1.86 (-6.65, 2.93) | -8.10 (-10.52, -5.68) | -8.75 (-11.27, -6.24) | -10.42 (-14.78, -6.07) |
| | Change in Tc | -6.37 (-8.83, -3.91) | -7.61 (-9.44, -5.77) | -8.45 (-10.29, -6.62) | -8.25 (-11.30, -5.20) |
| | Average Daily Tc | -6.69 (-8.89, -4.49) | -6.94 (-8.78, -5.10) | -7.95 (-10.03, -5.88) | -8.72 (-11.66, -5.77) |
| Cystatin-C eGFR (mL/min/1.73 m2) | Ambient Heat Index | 2.64 (-3.93, 9.21) | -1.04 (-5.66, 3.57) | -0.11 (-5.95, 5.73) | -1.20 (-8.98, 6.58) |
| | Change in Tc | 1.13 (-3.23, 5.49) | 0.31 (-2.65, 3.28) | -0.38 (-3.66, 2.90) | -1.24 (-5.90, 3.41) |
| | Average Daily Tc | 2.93 (-1.97, 7.84) | 2.11 (-1.09, 5.31) | -2.05 (-5.03, 0.94) | -2.96 (-7.90, 1.97) |
| Raw serum creatinine (mg/dL) | Ambient Heat Index | 0.05 (-0.02, 0.12) | 0.11 (0.07, 0.14) | 0.11 (0.07, 0.14) | 0.14 (0.08, 0.20) |
| | Change in Tc | 0.09 (0.06, 0.12) | 0.10 (0.08, 0.12) | 0.12 (0.09, 0.14) | 0.11 (0.08, 0.15) |
| | Average Daily Tc | 0.08 (0.06, 0.11) | 0.09 (0.06, 0.11) | 0.12 (0.09, 0.14) | 0.13 (0.09, 0.17) |
| Raw serum cystatin C (mg/L) | Ambient Heat Index | 0.10 (0.02, 0.18) | 0.14 (0.10, 0.19) | 0.13 (0.08, 0.18) | 0.13 (0.06, 0.21) |
| | Change in Tc | 0.12 (0.07, 0.17) | 0.13 (0.09, 0.17) | 0.13 (0.09, 0.17) | 0.13 (0.08, 0.18) |
| | Average Daily Tc | 0.12 (0.06, 0.17) | 0.11 (0.07, 0.15) | 0.15 (0.11, 0.19) | 0.14 (0.08, 0.20) |

Table S5. Doubly-robust modified exposure policy (MEP) estimates for the cross-shift change in the same four outcomes under a 2 °C additive reduction and a 5% multiplicative reduction in each heat metric, with 95% confidence intervals. Units and sign conventions follow Table S4.

| **Outcome** | **Exposure** | **Additive (-2 °C)** | **Multiplicative (-5%)** |
|---|---|---|---|
| Creatinine eGFR (mL/min/1.73 m2) | Ambient Heat Index | 3.00 (0.92, 5.09) | 2.94 (0.96, 4.91) |
| | Change in Tc | 2.47 (-2.60, 7.54) | 0.10 (-0.23, 0.44) |
| | Average Daily Tc | 1.58 (-3.54, 6.70) | 1.41 (-3.65, 6.47) |
| Cystatin-C eGFR (mL/min/1.73 m2) | Ambient Heat Index | 1.61 (-2.15, 5.37) | 1.28 (-2.34, 4.91) |
| | Change in Tc | 1.69 (-10.07, 13.45) | 0.17 (-0.34, 0.68) |
| | Average Daily Tc | 8.77 (-6.54, 24.08) | 8.29 (-5.96, 22.55) |
| Raw serum creatinine (mg/dL) | Ambient Heat Index | -0.03 (-0.06, 0.00) | -0.03 (-0.05, 0.00) |
| | Change in Tc | -0.04 (-0.10, 0.02) | 0.00 (-0.01, 0.00) |
| | Average Daily Tc | -0.07 (-0.14, 0.00) | -0.07 (-0.14, 0.01) |
| Raw serum cystatin C (mg/L) | Ambient Heat Index | -0.01 (-0.06, 0.03) | -0.01 (-0.06, 0.03) |
| | Change in Tc | -0.01 (-0.10, 0.07) | 0.00 (-0.01, 0.01) |
| | Average Daily Tc | -0.03 (-0.15, 0.09) | -0.03 (-0.16, 0.09) |
